# Elevated maternal hemoglobin levels during pregnancy are associated with autism spectrum disorder in the offspring: A population-based cohort study

**DOI:** 10.64898/2026.11.15.26368133

**Authors:** Amit Dichter, Alon Tzoref, Gal Meiri, Idan Menashe

**Author notes:** Contributed equally as co-first authors (Amit Dichter, Alon Tzoref). <u>**Contact Information**</u> Idan Menashe, PhD, Ben-Gurion University of the Negev, Beer-Sheva, Israel.

## Abstract

**Background:** Autism spectrum disorder (ASD) is an increasingly prevalent neurodevelopmental condition with multifactorial origins, including potential prenatal environmental contributions. Maternal anemia (decreased hemoglobin [Hb] levels) has long been associated with adverse offspring neurodevelopmental outcomes, yet evidence regarding elevated levels of antenatal hemoglobin levels, in relation to ASD risk remains limited. This study evaluated the association between maternal Hb levels during pregnancy and the risk of ASD diagnosis in offspring.

**Methods:** This large-scale, population-based retrospective cohort study used electronic health records from births of members of Clalit Health Services (CHS), the largest health maintenance organization in Israel, between 2011 and 2021. The study included 303,234 singleton live births to Jewish mothers who had at least one measured prenatal Hb value and were CHS members. Exclusion criteria were children with known genetic disorders and births <25 weeks of gestation. Maternal Hb, and red blood cells (RBC) levels during the first and second trimesters were categorized as low (<2.5th percentile), normal (reference), or high (>97.5th percentile). The main outcome was a diagnosis of ASD in the offspring, as recorded in medical files. Statistical analyses adjusted for maternal age, socioeconomic status, gestational diabetes, BMI, and other clinical and demographic factors.

**Results:** Among the 303,234 births, 8,106 (2.7%) children were diagnosed with ASD. High maternal Hb levels were associated with elevated odds of ASD diagnosis (aOR=1.22; 95% CI=1.08-1.37), with the strongest association observed for first-trimester measurements (aOR=1.31; 95% CI=1.14-1.49). This association persisted in the sibling-matched analysis (aOR=1.64; 95% CI=1.08–2.47) and was not modified by maternal RBC levels, suggesting that it was not explained by familial factors or RBC counts. The main limitations of this study include the lack of available data on maternal smoking, and the absence of detailed information on maternal nutritional status.

**Conclusions:** These findings suggest that elevated antenatal Hb levels may be relevant to offspring neurodevelopment. Further studies are needed to replicate these findings and clarify the biological mechanisms underlying this association.

## Introduction

Autism spectrum disorder (ASD) is a complex neurodevelopmental condition affecting 1-3% of children worldwide. [1] Over recent decades, ASD prevalence has continued to rise worldwide, a trend primarily attributed to evolving diagnostic practices and growing public awareness. However, the contribution of environmental factors to this increase in prevalence cannot be ruled out. [2–4].

The etiology of ASD is multifactorial, involving both genetic predispositions and diverse prenatal environmental influences. [2,5–7] Maternal health during pregnancy, including infections, use of medications, thyroid dysfunction, and hormonal variations, has increasingly been recognized as potentially impacting fetal neurodevelopment. [8–18] Beyond these factors, oxidative stress has emerged as a significant pathological mechanism that may further impair early brain development. This sustained imbalance between the production of reactive oxygen species and the body’s antioxidant defense systems is thought to induce molecular damage and exacerbate the clinical symptoms of the disorder.

Hemoglobin (Hb) plays a critical role in fetal oxygen delivery and is routinely measured, particularly during early gestation, as an essential component of prenatal care.[19] Abnormal maternal Hb levels may arise from various medical, nutritional, physiological, and environmental factors and may influence placental function and fetal brain development.[20,21] Low Hb levels (anemia) may reduce the oxygen-carrying capacity of maternal blood and contribute to fetal hypoxia, whereas elevated Hb levels may reflect inadequate plasma-volume expansion or hemoconcentration, potentially increasing blood viscosity and impairing uteroplacental perfusion. Thus, both low and high maternal Hb levels could compromise fetal oxygenation and create neurodevelopmental vulnerabilities relevant to autism spectrum disorder (ASD).[22–27]

Although several studies have examined the association between maternal anemia and ASD in offspring, their findings have been inconsistent, with some reporting increased risk only among children with co-occurring intellectual disability or under specific exposure conditions.[28–30] In contrast, the potential association between elevated maternal Hb levels and ASD has received little attention, despite evidence linking high Hb levels to adverse placental and pregnancy outcomes. Large-scale studies examining the full range and gestational patterns of maternal Hb levels are therefore needed to determine whether both low and high Hb levels are associated with ASD in offspring and to clarify the prenatal mechanisms that may underlie these associations.

This study aims to examine the association between maternal Hb concentrations during pregnancy, both below and above the normal range, and the subsequent risk of ASD in offspring. Clarifying these relationships will improve understanding of modifiable prenatal risk factors for ASD and may contribute to strategies for earlier diagnosis and intervention.

## Methods

### Study Design and Population

This retrospective cohort study utilized electronic health records from Clalit Health Services (CHS), Israel’s largest healthcare provider. The study population included all singleton live births > 25 weeks of gestation of CSH members, between 2011 and 2021. The analysis was restricted to births of Jewish mothers due to >20% of missing gestational complete blood count (CBC) measures in other sub-populations due to suboptimal utilization of routine prenatal care [31]. We excluded children with known genetic syndromes associated with ASD[30], and absence of maternal Hb level measurements during the first or second trimester. Consequently, the study sample included 303,234 children. Of these, 8,106 children (2.7%) were diagnosed with ASD, which is consistent with national estimates[4]. In addition, a nested matched-sibling analysis was conducted, including 2,894 children diagnosed with ASD and one of their non-ASD siblings. A flowchart with the study sample ascertainment is depicted in **Figure 1**.

**Figure 1:**
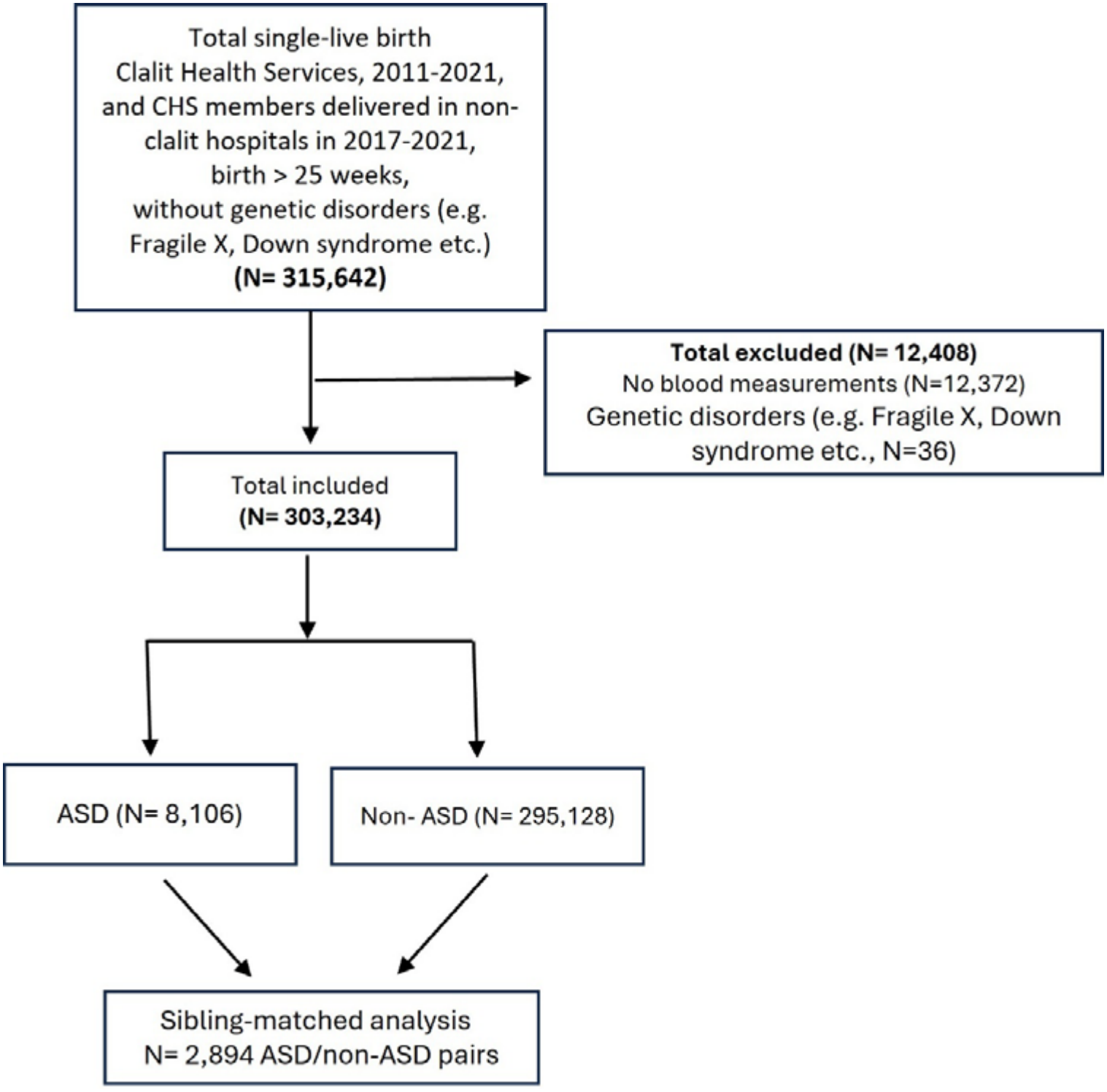
Flowchart of cohort ascertainment.

### Data Collection

Data collection incorporated a broad array of maternal and child characteristics, laboratory test results, and clinical outcomes obtained from the CSH electronic database via the MDclone system and subsequently analyzed in the virtual desktop infrastructure (VDI) environment of CHS. To ensure the integrity and validity of the statistical analyses, only variables with less than 20% missing data were included. Maternal Hb, red blood cells (RBC) levels were obtained from routine CBC tests performed in the first and/or second trimester of pregnancy. Laboratory values were rigorously filtered according to physiologically reasonable limits, with maternal Hb restricted to the range of 4–20 g/dL in line with established literature. [32]

### Exposure Assessment

We classified pregnancies according to their maternal Hb, and RBC measures into “low” (<2.5^th^ percentile), or “high” (>97.5^th^ percentile) groups in either the first or second trimesters, with the middle 95% serving as the “normal” reference group. Trimester-specific analyses were also conducted to evaluate the exposures from each period independently.

### Outcome Definition

The primary outcome for the study was a documented diagnosis of ASD in offspring (ICD-9 code 299.0, autistic disorder) as recorded in the CHS database. In Israel, ASD diagnosis requires both medical and psychological assessments. Because documentation of an ASD diagnosis, according to DSM-5 criteria [5] is a prerequisite for state-funded benefits, including ASD-related health services, it is assumed that most children who are members of CHS with a formal diagnosis of ASD are documented as such in the CHS database.

### Covariates

Covariates included maternal age, socioeconomic status, offspring sex, maternal BMI, gestational diabetes, gestational hypertension, birth weight, and gestational age. All included variables satisfied the criterion of less than 20% missingness.

### Statistical Analysis

Standard univariate analyses were used to compare maternal and neonatal characteristics between the study groups. Multivariable logistic regression models were then used to assess the associations of low and high maternal hemoglobin (Hb) levels, relative to the normal range, with the odds of ASD in offspring. As a secondary analysis, we examined the associations of low and high maternal red blood cell (RBC) counts with offspring ASD. All models were adjusted for relevant sociodemographic and clinical confounders. To account for shared familial and genetic factors, we also conducted a sibling-matched analysis using conditional logistic regression, in which each ASD case was matched to one randomly selected non-ASD sibling. The Breslow–Day test was used to assess the homogeneity of odds ratios (ORs) across stratified subsamples.

All statistical analyses were performed using R software (version 4.2.2), and statistical significance was set at a p-value < 0.05.

### Ethical Approval

The study was approved by the Institutional Review Board of Soroka Medical Center (SOR-237-21).

## Results

A total of 303,234 singleton live births were included in the study sample (**Figure 1**). The distributions of Hb levels approximated normal curves and were significantly correlated across trimesters (**Figure 2**). As expected, maternal Hb levels have slightly decreased from 1^st^ to 2^nd^ trimester (Mean Hb: 12.52±0.94 g/dL to 11.56±0.91; p <.001), probably due to increased plasma volume during pregnancy. The corresponding lowest 2.5^th^ and highest 97.5^th^ percentiles Hb level cutoffs were ≤10.6 g/dL and high Hb as ≥14.3 g/dL for the first trimester and ≤9.7 g/dL and ≥13.3 g/dL for the second trimester. RBC counts showed similar distributions and inter-trimester correlations (**Figure S1**).

**Figure 2.**
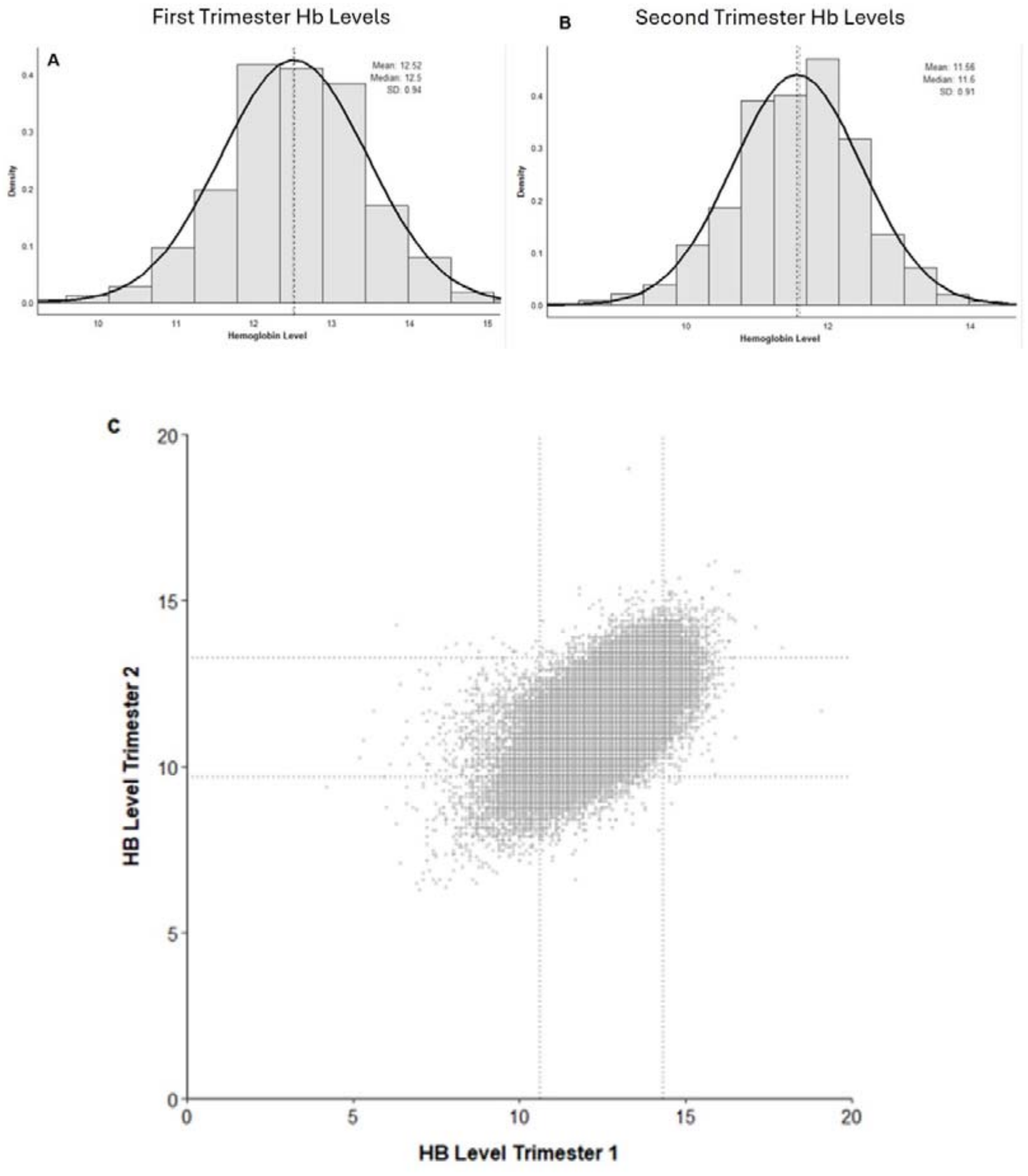
Distribution of Hb levels during pregnancy. Histograms representing maternal Hb levels during the first (A) and second (B) trimesters. The Hb distribution at both trimesters is approximately normal with a mean Hb level of 12.52 g/dL (SD 0.94) in the first trimester and 11.56 g/dL (SD 0.91) at the second trimester. A scatter plot (C) depicting a significant positive correlation between first and second maternal Hb levels (Pearson r = 0.61, P < .001). Dashed horizontal and vertical lines represent the 2.5th and 97.5th percentile cutoff values used to define the study groups. In the first trimester, these cutoffs were ≤10.6 g/dL and ≥14.3 g/dL; in the second trimester, corresponding values were ≤9.7 g/dL and ≥13.3 g/dL.

Table 1 presents maternal and neonatal characteristics across Hb groups. While many group differences are statistically significant due to the large sample size, the absolute differences are generally small. The only notable difference is the slightly higher rates of gestational diabetes and hypertension among women with elevated Hb levels. Notably, the prevalence of ASD diagnosis was significantly higher in the high Hb group compared to the normal Hb group (3.3% vs. 2.6%; p<0.001), while the low Hb group (2.8%) did not show such difference.

In the multivariable regression models (**Table 2**), high maternal Hb levels during either 1^st^ or 2^nd^ trimesters were associated with increased odds of ASD in the offspring (aOR=1.22; 95% CI: 1.08-1.37) with slightly higher OR seen in the 1^st^ trimester (aOR=1.31; 95% CI: 1.14-1.49) than in the 2^nd^ trimester (aOR=1.15; 95% CI: 1.01-1.33). Conversely, low Hb levels were not associated with ASD nor any of the extreme RBC levels (**Table 2**). Similar results were observed when the cutoffs for the extreme groups were extended to the 5^th^ and 95^th^ percentiles (Table S1), except for a moderate association of low Hb levels during the 2^nd^ trimester with decreased ASD odds (aOR=0.85; 95% CI: 0.75-0.97).

**Table 1:** Characteristics of the study population across the study groups.

| **Variable** |  | **Maternal Hb levels** |  |  |  |  |
|---|---|---|---|---|---|---|
|  |  | **Normal (Ref)** <sup>a</sup><br>**(N = 283,365)** | **Low** <sup>b</sup><br>**(N=9,614)** | **High** <sup>c</sup><br>**(N=10,255)** | **P-Value**<br>**(a vs. b)** | **P-Value**<br>**(a vs. c)** |
| **Maternal age**<br>(mean±SD) |  | 31.6 (±5.1) | 31.9 (±5.5) | 31.1 (±5.1) | **<0.001** | **<0.001** |
| **Offspring sex**<br>[N(%)] | **Male** | 145,514 (51%) | 4,790 (50%) | 5,408 (53%) | **0.003** | **0.006** |
|  | **Female** | 137,851 (49%) | 4,824 (50%) | 4,847 (47%) |  |  |
| **Socioeconomic status**<br>[N(%)] | **Very low** | 25,207 (8.1%) | 1,563 (18%) | 126 (1.3%) | **<0.001** | **<0.001** |
|  | **Low** | 54,702 (20%) | 2,408 (27%) | 1,962 (21%) |  |  |
|  | **Medium** | 99,152 (37%) | 3,768 (42%) | 3,716 (39%) |  |  |
|  | **High** | 85,134 (32%) | 2,148 (24%) | 2,861 (30%) |  |  |
|  | **Very high** | 24,546 (9.2%) | 484 (5.4%) | 855 (9%) |  |  |
| **Maternal BMI**<br>[mean (± SD)] |  | 22.1 (±4.3) | 22.6 (±5.5) | 22.4 (±4.4) | **<0.001** | **<0.001** |
| **Gestational Diabetes**<br>[N(%)] |  | 15,072 (5.3%) | 479 (5%) | 744 (7.3%) | 0.15 | **<0.001** |
| **Gestational Hypertension**<br>[N(%)] |  | 9,917 (3.5%) | 369 (3.8%) | 606 (5.9%) | 0.076 | **<0.001** |
| **IVF**<br>[N(%)] |  | 10,738 (3.8%) | 410 (4.3%) | 385 (3.8%) | **0.017** | 0.9 |
| **Birth weight (gr)**<br>[mean (± SD)] |  | 3,286 (±433) | 3,251 (±454) | 3,247 (±450) | **<0.001** | **<0.001** |
| **Birth Week**<br>[mean (± SD)] |  | 39.15 (±1.40) | 39.02 (±1.52) | 39.08 (±1.51) | **<0.001** | **<0.001** |
| **ASD Diagnosis**<br>[N(%)] |  | 7,497 (2.6%) | 270 (2.8%) | 339 (3.3%) | 0.3 | **<0.001** |

**Table 2:** Results of logistic regression models for the association between low/high Hb levels and ASD.

| **Variable** |  | **Trimester** | **Crude analysis** |  | **Adjusted analysis\*** |  |
|---|---|---|---|---|---|---|
|  |  |  | **OR (95% CI)** | **P-Value** | **OR (95% CI)** | **P-Value** |
| **Hb levels** | **Low** | **1<sup>st</sup> or 2<sup>nd</sup>** | 1.06 (0.94-1.2) | 0.329 | 1.02 (0.89-1.16) | 0.754 |
|  |  | **1<sup>st</sup>** | 1.07 (0.92-1.23) | 0.35 | 1.02 (0.88-1.19) | 0.747 |
|  |  | **2<sup>nd</sup>** | 0.95 (0.82-1.1) | 0.55 | 0.93 (0.79-1.09) | 0.408 |
|  | **High** | **1<sup>st</sup> or 2<sup>nd</sup>** | **1.26 (1.12-1.4)** | **<0.001** | **1.22 (1.08-1.37)** | **<0.001** |
|  |  | **1<sup>st</sup>** | **1.32 (1.16-1.49)** | **<0.001** | **1.31 (1.14-1.49)** | **<0.001** |
|  |  | **2<sup>nd</sup>** | **1.18 (1.02-1.34)** | **0.01** | **1.15 (1.01-1.33)** | **0.04** |
| **RBC levels** | **Low** | **1<sup>st</sup> or 2<sup>nd</sup>** | 0.89 (0.78-1.01) | 0.082 | 0.95 (0.83-1.08) | 0.464 |
|  |  | **1<sup>st</sup>** | 0.90 (0.79-1.05) | 0.185 | 0.92 (0.78-1.08) | 0.353 |
|  |  | **2<sup>nd</sup>** | 0.91 (0.77-1.07) | 0.258 | 1.0 (0.84-1.19) | 0.925 |
|  | **High** | **1<sup>st</sup> or 2<sup>nd</sup>** | **1.16 (1.03-1.29)** | **0.011** | 1.06 (0.94-1.19) | 0.35 |
|  |  | **1<sup>st</sup>** | 1.11 (0.96-1.27) | 0.168 | 1.02 (0.87-1.18) | 0.824 |
|  |  | **2<sup>nd</sup>** | **1.18 (1.02-1.35)** | **0.026** | 1.07 (0.91-1.24) | 0.395 |

\*Adjusted for the following socio-economic and clinical variables: (Socio-economic status, child gender, maternal age, maternal BMI, maternal diabetes, maternal hypertension, birth weight, birth week)

### Matched-Sibling Analysis

Repeating the regression analyses in the matched-sibling sample reproduced similar results as in the main analysis (**Table 3**). Specifically, high maternal Hb levels were associated with increased ASD risk within families (adjusted OR=1.64; 95% CI: 1.08-2.47) whereas low Hb and either high or low RBC were not associated with ASD in these families.

**Table 3:**
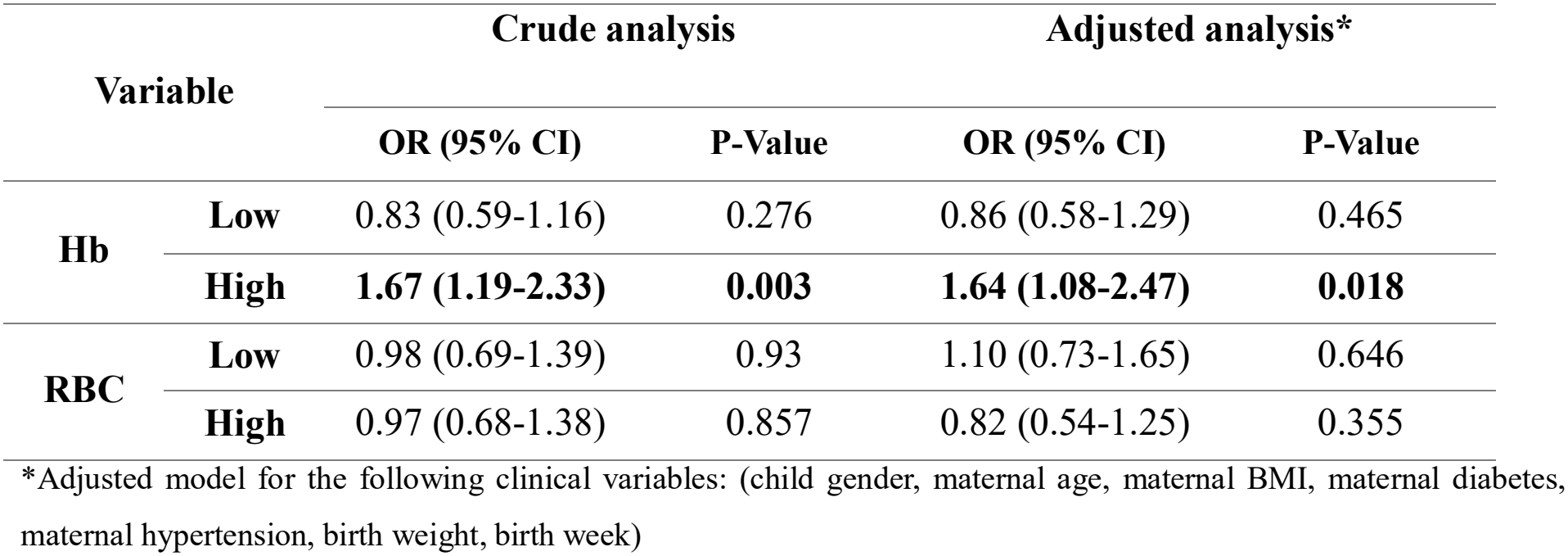
Results of the matched-sibling analysis.

### Stratification Analysis

Finally, to examine whether RBC count contributed to the observed association between elevated maternal Hb levels and ASD in offspring, we repeated the primary regression analysis in a stratified cohort by the median of maternal RBC count. The results are presented in supplementary **Table S2**. In the full-cohort analysis, similar adjusted ORs were seen in the two RBC strata (adjusted OR, 1.26; 95% CI, 1.01-1.54 and adjusted OR, 1.28; 95% CI, 1.03-1.58 for RBC≤50% and RBC>50% respectively; Breslow-Day p-value = 0.353). In the matched-sibling analysis, the adjusted OR was higher among pregnancies with RBC counts at or below the median than among those with RBC counts above the median (adjusted OR, 2.28; 95% CI, 1.03–5.20 vs adjusted OR, 1.12; 95% CI, 0.60–2.08). However, this difference was not statistically significant (Breslow–Day p = 0.110). These findings suggest that maternal RBC count does not modify the association between elevated maternal Hb levels and ASD in offspring, although the difference in point estimates in the sibling-matched analysis warrants cautious interpretation.

## Discussion

This is one of the largest and most comprehensive population-based cohort study to examine the association between abnormal antenatal Hb levels and ASD in offspring. Our findings indicate that elevated maternal Hb levels during the first or second trimester are associated with increased odds of ASD in offspring and that this association is unlikely to be explained by shared familial factors or maternal RBC levels.

To our knowledge, this is the first study to systematically identify an association between elevated maternal Hb levels and ASD in offspring. In contrast, we found no association between low maternal Hb levels and ASD, consistent with two previous studies,[29,30] although another study reported an association between anemia in early pregnancy and increased ASD risk.[28] Together, these findings extend the literature beyond maternal anemia and suggest that elevated antenatal Hb may reflect prenatal physiological processes relevant to fetal neurodevelopment.

The association between elevated maternal Hb levels and ASD in offspring remained consistent across multiple definitions of high Hb and was not modified by maternal RBC counts. Moreover, the association persisted in the sibling-matched analysis, which accounted for shared familial factors. Together, these convergent findings strengthen the robustness of the observed association and suggest that it may reflect pregnancy-specific biological processes rather than confounding by familial or other factors.

Elevated Hb levels may reflect impaired plasma volume expansion during pregnancy, increased blood viscosity, reduced placental perfusion, or decreased nitric oxide bioavailability,[33,34] all of which could compromise fetal oxygenation and affect neurodevelopment. Alternatively, elevated Hb may indicate increased iron stores, as excess iron can promote oxidative stress and adverse placental processes.[35] The finding that the association between elevated maternal Hb and offspring ASD was not modified by RBC levels may be more consistent with Hb-related mechanisms, such as altered iron homeostasis or oxidative stress, than with mechanisms driven primarily by RBC counts. However, it does not exclude potential contributions from increased blood viscosity or impaired placental perfusion. Hence, the specific mechanisms underlying this association warrant further investigation

### Strengths and Limitations

The strengths of this study include its large, well-characterized cohort; prospectively recorded laboratory and clinical data; repeated measures across two trimesters; adjustment for various sociodemographic, maternal and perinatal variable, as well as for familial confounding via the matched-sibling analysis. However, it also had several limitations that warrant further consideration. First, the restriction of our cohort only to Jewish mothers, may limit the generalizability of our findings to other ethnic populations. Second, the use of routinely collected electronic health records may include few data entry errors or misclassification. However, the large sample size and the use of physiologically validated ranges for laboratory values likely mitigate the impact of such inaccuracies. Third, data on maternal smoking, a known correlate of Hb levels, was unavailable in the CHS database, precluding our ability to adjust for this specific potential confounder. Lastly, information on factors that may influence maternal Hb levels during pregnancy, including nutritional status, iron supplementation, hydration status, and other health conditions was unavailable, limiting our ability to further explore the mechanisms underlying the observed association.

### Conclusions

Elevated maternal Hb levels during early and mid-pregnancy were consistently associated with increased odds of ASD in offspring across multiple analytic approaches. These findings identify elevated antenatal Hb as a potentially underrecognized indicator of prenatal processes relevant to neurodevelopment and warrant further investigation of the underlying biological mechanisms.

## Data Availability

Raw data from this study could not be shared due to institutional restrictions. Summary data and statistics produced in the present study are available upon reasonable request to the authors

