## Supplementary material for "Elevated maternal hemoglobin levels during pregnancy are associated with autism spectrum disorder in the offspring: A population-based cohort study": Table S1

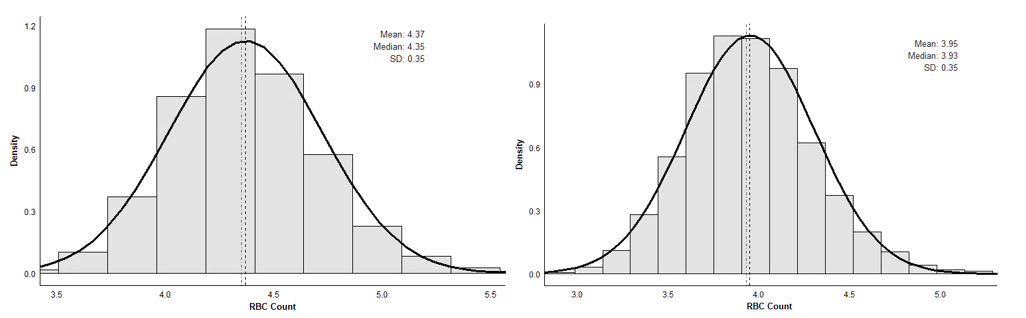
**Figure S1: Distribution of RBC levels during pregnancy**

Second Trimester RBC Levels

First Trimester RBC Levels

**A**

**B**

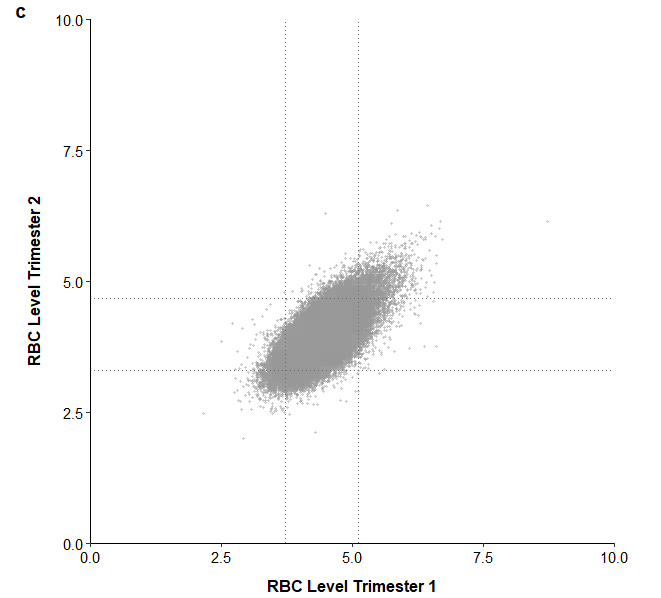

**Figure S1. Distribution of RBC levels during pregnancy**

Histograms representing maternal RBC counts during the first (A) and second (B) trimesters. The RBC distribution at both trimesters is approximately normal with a mean RBC count of 4.37 ×10^6^/µL (SD 0.35) in the first trimester and 3.95 ×10^6^/µL (SD 0.35) in the second trimester. A scatter plot (C) depicting a significant positive correlation between first and second maternal RBC counts (Pearson r = 0.68, *P* < .001). Dashed horizontal and vertical lines represent the 2.5^th^ and 97.5^th^ percentile cutoff values used to define the study groups. In the first trimester, these cutoffs were ≤3.72 × 10⁶/µL and ≥5.11 × 10⁶/µL. In the second trimester, corresponding values were ≤3.30 × 10⁶/µL and ≥4.69 × 10⁶/µL.

**Table S1: Logistic regression models – 5% cutoff**

| Variable | | Trimester | | Crude analysis | | Adjusted analysis* | |
| --- | --- | --- | --- | --- | --- | --- | --- |
|  |  |  |  | **OR (95% CI)** | **P-Value** | **OR (95% CI)** | **P-Value** |
| **Hb levels** | **Low** | | **1^st^ or 2^nd^** | 0.99 (0.91-1.09) | 0.985 | 0.97 (0.88-1.06) | 0.558 |
|  |  |  | **1^st^** | 1.02 (0.91-1.14) | 0.662 | 1.00 (0.89-1.12) | 0.929 |
|  |  |  | **2^nd^** | **0.87 (0.77-0.98)** | **0.028** | **0.85 (0.75-0.97)** | **0.019** |
|  | **High** | | **1^st^ or 2^nd^** | **1.22 (1.12-1.32)** | **<0.001** | **1.19 (1.09-1.29)** | **<0.001** |
|  |  |  | **1^st^** | **1.26 (1.13-1.39)** | **<0.001** | **1.25 (1.12-1.39)** | **<0.001** |
|  |  |  | **2^nd^** | **1.17 (1.05-1.31)** | **0.003** | **1.14 (1.01-1.27)** | **0.023** |
| **RBC levels** | **Low** | | **1^st^ or 2^nd^** | 0.91 (0.83-1.00) | 0.053 | 0.96 (0.87-1.05) | 0.459 |
|  |  |  | **1^st^** | 0.93 (0.83-1.04) | 0.221 | 0.96 (0.85-1.08) | 0.568 |
|  |  |  | **2^nd^** | **0.86 (0.76-0.97)** | **0.022** | 1.0 (0.84-1.19) | 0.343 |
|  | **High** | | **1^st^ or 2^nd^** | **1.14 (1.05-1.24)** | **0.001** | 1.04 (0.95-1.14) | 0.302 |
|  |  |  | **1^st^** | **1.13 (1.02-1.25)** | **0.014** | 1.06 (0.94-1.18) | 0.283 |
|  |  |  | **2^nd^** | **1.14 (1.02-1.26)** | **0.016** | 1.01 (0.90-1.13) | 0.769 |

*Adjusted for the following socio-economic and clinical variables: (Socio-economic status, child gender, maternal age, maternal BMI, maternal diabetes, maternal hypertension, birth weight, birth week)

**Table S2. Stratification analysis**

| Variable | | Adjusted OR (95% CI) * | | Breslow-Day test of homogeneity |
| --- | --- | --- | --- | --- |
|  |  | **RBC ≤ 50%** | **RBC > 50%** |  |
| High Hb (>97.5%) | **Full Cohort** | 1.26 (1.01-1.54) | 1.28 (1.03-1.58) | p=0.353 |
|  | **Matched-siblings** | 2.28 (1.03-5.20) | 1.12 (0.60-2.08) | p=0.110 |

*Adjusted model for the following clinical variables: (child gender, maternal age, maternal BMI, maternal diabetes, maternal hypertension, birth weight, birth week)
